# Evaluating an Ambient Artificial Intelligence Scribe for Documentation Quality and Efficiency in Psychiatric Consultations: A Simulation-based Study

**DOI:** 10.1101/2025.09.21.25336260

**Authors:** Faisal A. Nawaz, Syed Ali Bokhari, Firdous M. Usman, Zara Arshad, Meghana Sudhir, Ralf Krage, Rahul Kashyap

**Affiliations:** Al Amal Psychiatric Hospital, Emirates Health Services, Dubai, United Arab Emirates; Global Remote Research Scholars’ Program, Princeton Junction, NJ, United States; American Hospital Dubai, Dubai, United Arab Emirates; Institute of Learning, Mohammed Bin Rashid University of Medicine and Health Sciences, Dubai, United Arab Emirates; Department of Research, WellSpan Health, York, Pennsylvania, United States

**Keywords:** ambient AI, artificial intelligence, ambient scribes, documentation, informatics, electronic health records, large language models, medical scribe

## Abstract

**Background:** Documentation demands in psychiatric practice diminish time for direct patient care and are associated with clinician burnout. Ambient artificial intelligence (AI) scribes may facilitate more efficient and higher-quality documentation while reducing clinician workload and preserving the integrity of the clinical encounter. This study aims to determine the impact of an ambient AI scribe in improving documentation quality and efficiency while reducing clinician workload during simulated psychiatric consultations.

**Methods:** We conducted a prospective, cross-over, within-subject simulation study to compare conventional keyboard-based documentation (*traditional condition*) with documentation assisted by an ambient artificial-intelligence scribe (*AI-scribe condition*). The study was conducted at an academic simulation centre comprising of eight clinicians with psychiatric experience who completed both documentation conditions. Clinician workload (NASA Task Load Index [NASA-TLX]), documentation quality (Sheffield Assessment Instrument for Letters [SAIL]), video-verified screen time during documentation, and bespoke clinician and patient experience questionnaires were administered.

**Results:** AI-scribe use was associated with substantially lower workload versus traditional documentation (NASA-TLX total, 25.0 versus 461.3; mean difference, 436.25; 95% CI, 389.93-482.57; p<0.001), with significant improvements in 5 of 6 subscales, including temporal demand (mean difference, 50.00; p<0.001), frustration (mean difference, 33.75; p=0.04), and perceived performance (mean difference, 30.0; p=0.03). Documentation quality improved with AI-scribe (SAIL; 22.88 versus 14.38; t(7)=2.55; p=0.038; Cohen’s dz=0.90); checklist completeness was ≥75% on 17 of 20 items, with 9 items at 100%. Screen time decreased (7.46 versus 5.31 minutes; mean difference, −2.15; 95% CI, −2.10-+6.41; p=0.27; Hedges g=0.38); 5 of 8 clinicians showed individual reductions. Overall clinician satisfaction was higher with AI-scribe (100% versus 50%), and 87.5% agreed that AI assistance reduced cognitive load. Patient-reported experience favoured the AI-scribe condition.

**Conclusion:** Ambient AI scribe can assist in improving documentation quality and substantially reducing clinician workload while maintaining favourable patient-perceived consultation quality.

## Introduction

Since the introduction of Electronic Health Records (EHR) in the 1960s, there has been a considerable shift in the approach to patient care (Ali et al., 2023). Clinicians are expected to spend increased time and effort documenting patient data and balancing patient interactions during consultations. Studies report that clinicians dedicate around 35% of their time to documentation rather than direct patient care (Joukes et al., 2018). Psychiatrists are particularly vulnerable to increased workload burden, with studies showing on average three hours per work day spent in documentation (Spießl and Hausner, 2012).

Currently, this approach reduces the overall time spent on patient interactions and contributes to the rising burnout rates in the medical field (Mache et al., 2011). To tackle this challenge, ambient Artificial Intelligence (AI) scribes have shown potential by automating note-taking in healthcare settings using Natural Language Processing (NLP) and Large Language Models (LLM) (Mess et al., 2025). The technology relies on listening to conversations during consultations and identifying the relevant clinical information to be extracted into an editable documentation format. Studies evaluating AI scribes have shown improvement in documentation quality, efficiency, and overall job satisfaction among healthcare professionals (Balloch et al., 2024; Doshi et al., 2024; Lee et al., 2024).To date, there are no studies that have evaluated its impact among psychiatrists. Our pilot study aims to evaluate the documentation quality, efficiency, and experiences of psychiatrists utilizing AI scribes in a simulated setting.

## Methods

### Study design and setting

We conducted a prospective, cross-over, within-subject simulation study to compare conventional keyboard-based documentation (*traditional condition*) with documentation assisted by an ambient artificial-intelligence scribe (*AI-scribe condition*). The study was conducted at an academic simulation centre comprising eight consultation rooms. Participants utilized their personal laptops, each connected to a standardized simulated electronic health record (EHR) environment. The sequence of consultation group sessions is illustrated as a flow diagram in **Figure 1**.

**Figure 1.**
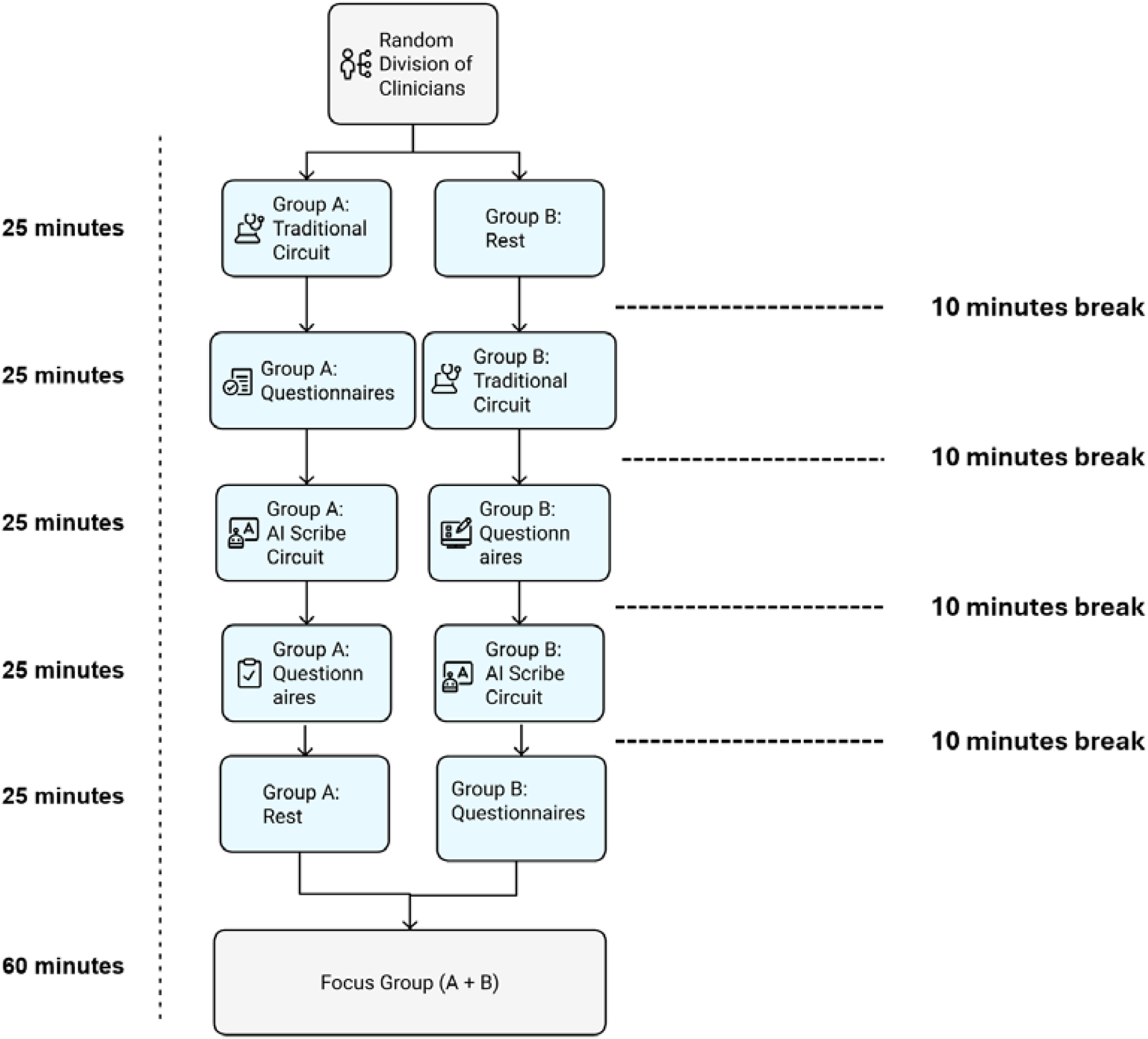
Outline of Simulation Circuit for Groups A and B.

### Participants

#### Recruitment and eligibility

Eight clinicians from a tertiary psychiatric hospital in Dubai, United Arab Emirates, were invited to participate, comprising three consultants, two specialists, and three residents. The inclusion criteria required a minimum of two years of clinical experience.

#### Pre-study consent and confidentiality

Prior to commencement of the data collection, participants and simulated patients (SPs) received and agreed to the confidentiality agreement with a written consent form to participate in the study.

#### Ambient AI-scribe intervention

The ambient AI-scribe system continuously captured clinician-patient dialogue through an omni-directional microphone. It transcribed the speech into text and extracted clinically relevant information using a rules-based chunker. This system automatically labelled speakers, identified key topic sentences, and mapped each to the corresponding Systematized Nomenclature of Medicine Clinical Terms (SNOMED CT) concept. By employing natural language processing and sophisticated language model algorithms, the AI scribe generated an editable draft consultation note directly within its interface. Clinicians were permitted to add or delete text within the AI-generated draft; however, they were instructed not to rewrite the entire consultation note.

Prior to each consultation, clinicians selected a standardized psychiatric documentation template available within the AI-scribe platform. This template was modified for the study to ensure consistency in documentation structure, clinical comprehensiveness, and alignment with the bio-psycho-social model of psychiatric formulation.

### Standardised patient scenarios

#### Case development

Eight adult outpatient scenarios targeting common outpatient clinical presentations were prepared in advance with the consensus of two external psychiatrists. They were suggestive of Bipolar disorder, Schizophrenia, Obsessive Compulsive Disorder, and Major Depressive Disorder.

### Actor recruitment and training

Four standardized SPs, who were experienced in psychiatric patient portrayal, participated in this study. They underwent training using the pre-planned scripts based on eight psychiatric case scenarios as above.

### Simulation circuit and randomisation

#### Room layout

Four identical rooms were arranged with a clinician desk and three chairs, one each for the clinician, patient, and an observer (included for quality assurance).

#### Randomisation procedure

Participants were randomized using a computer-generated sequence with a block size of four into two equal groups (Group A and Group B). Allocation was blinded to participants and standardised patients to avoid selection bias. In the first round, Group A completed four consultations using traditional documentation methods, followed by Group B completing four traditional consultations in the second round. Subsequently, during the third and fourth rounds, Group A and Group B completed four consultations each utilizing the AI-scribe method, in the same sequence. A 10-minute buffer separated each round to mitigate fatigue. An outline of the simulation circuit can be seen below (**Figure 1**).

### Data capture and Questionnaire Development

(Details in Supplement file A) Independent observers in each consultation room recorded Consultation duration and the proportion of eye contact directed toward the patients. Immediately following each station, clinicians were asked to complete the National Aeronautics and Space Administration Task Load Index (NASA-TLX), Clinician Questionnaire (CQ), and Experience Questionnaire (PEQ) (Hart and Staveland, 1988; Mercer et al., 2004; “Visit-Specific Satisfaction Instrument (VSQ-9),” n.d.).

To assess clinician experiences with both traditional and AI-assisted documentation, a clinician experience questionnaire was developed. It consisted of questions from Physician Worklife Survey (PWS) (McMurray et al., 1997), System Usability Scale (SUS) NASA-TLX (Hart and Staveland, 1988), the Electronic Health Record Nurse Satisfaction (EHRNS) Instrument(Sockolow et al., 2012) and the Mini-Z Burnout Survey.(Linzer et al., 2022) A patient experience questionnaire was used based on the Consultation and Relational Empathy (CARE) Measure, along with the Visit-Specific Satisfaction Questionnaire (VSQ-9) (Marshall and Hays, 1994; Mercer et al., 2004; “Visit-Specific Satisfaction Instrument (VSQ-9),” n.d.).

### Outcome measures

Four primary outcome measures were evaluated (**eTable 1**); Clinician workload by NASA-TLX(Hart and Staveland, 1988), documentation quality by the Sheffield Assessment Instrument for Letters (SAIL) (Crossley et al., 2001), Clinician experience by CQ and the SPs experience by Patient Experience Questionnaire (PEQ).

**Table 1.**
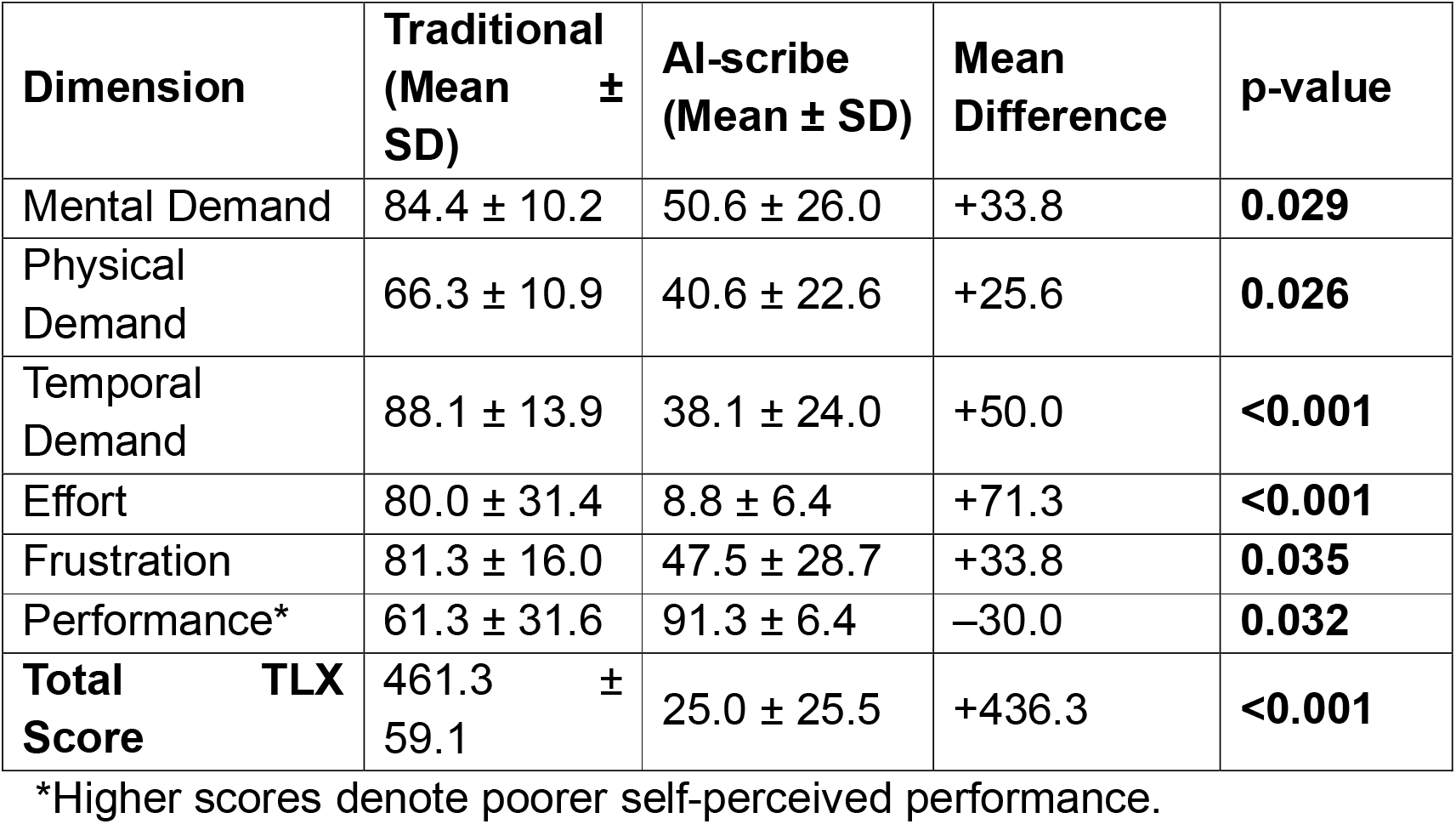
*NASA-TLX Scores by Documentation Method*. Positive mean differences (A – B) denote greater workload under the traditional method; higher scores indicate heavier perceived load.

### Data management

Paper instruments were double-entered into Microsoft Excel which was subsequently imported to SPSS v29.0.2.0. Electronic data (EHR notes, timing logs) were stored on an encrypted institutional server.

### Statistical analysis

Descriptive statistics were utilised for both continuous and categorical variables. For continuous variables, mean ± standard deviation (SD), median, and range were reported, while counts and percentages were provided for ordinal or binary variables. The primary analysis focused on within-clinician differences in NASA-TLX total scores between documentation methods, assessed using paired-samples t-tests (two-tailed), with effect sizes calculated using Hedges’ d to account for small sample corrections.

Paired-samples t-tests were performed for NASA-TLX sub-scales, SAIL totals, consultation time, and documentation time. McNemar tests were used to evaluate dichotomous SAIL items, while exact Pearson χ^2^ or Fisher’s tests assessed associations between different variables within each condition. No missing data were observed in the study. All inferential statistical tests were executed using SPSS software, and graphical representations were created with Microsoft Excel. Statistical significance was defined as p<0.05 (two-tailed) for both primary and secondary outcomes, and p-values were accompanied by 95% confidence intervals where informative. A power analysis (G*Power v3.1) showed that the sample of eight paired observations provided 91% power at α=0.05 (two-tailed) to detect the observed NASA-TLX total-score difference.

### Ethical considerations

The authors assert that all procedures contributing to this work comply with the ethical standards of the relevant national and institutional committees on human experimentation and with the Helsinki Declaration of 1975, as revised in 2013. All procedures involving human subjects/patients were approved by the Dubai Scientific Research Ethics Committee (DSREC) waived formal review (MBRU IRB-2025-203).

## Results

### Participant Characteristics and Data Completeness

All eight clinicians successfully completed assigned documentation tasks. The cohort consisted of four females and four males. Each clinician completed documentation twice: once using conventional typing (traditional method) and once with an ambient AI-scribe. A total of sixteen documentation yielded a total of 128 observations across all outcome measures.

### Clinician Workload (NASA-TLX)

Under the traditional documentation method, clinicians reported a high perceived workload burden (mean NASA-TLX score=461.3, SD=59.1; **Table 2**). In contrast, documentation assisted by AI-scribe resulted in a markedly reduced workload (mean=25.00, SD=25.49). A paired-samples t-test confirmed a statistically significant reduction in total workload (mean difference=436.25; 95% CI: 389.93-482.57).

**Table 2.** *SAIL Scores by Documentation Method*. For each participant, traditional and AI-scribe scores and their quality bands are listed; ΔSAIL is defined as AI-scribe minus traditional.

| **Participant** | **Traditional Score** | **Traditional Quality Band** | **AI-Scribe Score** | **AI-Scribe Quality Band** | **ΔSAIL** |
| --- | --- | --- | --- | --- | --- |
| P01 | 10 | Very Poor | 26 | Good | +16 |
| P02 | 17 | Very Poor | 25 | Good | +8 |
| P03 | 14 | Very Poor | 23 | Fair | +9 |
| P04 | 12 | Very Poor | 28 | Very Good | +16 |
| P05 | 19 | Poor | 21 | Poor | +2 |
| P06 | 15 | Very Poor | 22 | Fair | +7 |
| P07 | 13 | Very Poor | 24 | Fair | +11 |
| P08 | 15 | Very Poor | 29 | Very Good | +14 |

\*Positive values denote higher quality with the AI-scribe.

Significant improvements were observed across five of the six NASA-TLX domains, with the largest reductions in *Effort* (mean difference=71.25; 95% CI: 43.19-99.31) and *Temporal Demand* (mean difference=50.00; 95% CI: 32.13-67.88), indicating clinicians expended substantially less energy and felt notably less pressured when using the AI-scribe. Additionally, scores significantly decreased in *Mental Demand* (mean difference=33.75; p=0.03; 95% CI: 4.55-62.95), *Physical Demand* (mean difference=25.63; p=0.03; 95% CI: 4.14-47.11), and *Frustration* (mean difference = 33.75; p=0.04; 95% CI: 3.13-64.37), demonstrating reductions in cognitive, physical, and emotional burdens, respectively. Clinicians also perceived significantly improved *Performance*, indicating higher self-rated success in documentation tasks when using the AI-scribe (mean difference=30.00; t(7)=2.66; p=0.03; 95% CI: 3.38-56.63). Collectively, these findings demonstrate that AI-assisted documentation considerably reduces clinicians’ perceived workload while simultaneously enhancing their subjective performance.

### Documentation Quality (SAIL)

A paired-samples t-test was used to compare the two documentation methods (t(7)=2.55, p=0.038, Cohen’s dz=0.90) (**Table 3**). Documentation produced using the traditional method yielded a mean (±SD) SAIL total score of 14.38 (±6.14). This average score places the quality of traditionally generated notes predominantly within the “Very Poor”-”Poor” categories, highlighting significant deficiencies. In contrast, the AI-scribe generated notes achieved a significantly higher mean of 22.88 (±4.32). This score situates the AI-scribe notes primarily in the “Fair”-”Good” range. AI-scribe documentation demonstrated higher compliance with content expectations where 17 of 20 checklist items showed greater than or equal to 75% completion, with nine items achieving 100% completion.

**Table 3.** *Clinician Screen*⍰*Time by Documentation Method*. Δ Screen-time is defined as the difference between AI-scribe and traditional conditions.

| Participant | Traditional Screen Time (mins) | AI Scribe Screen Time (mins) | $\Delta$ Screen Time (mins)* |
| --- | --- | --- | --- |
| P01 | 4.57 | 8.20 | -3.63 |
| P02 | 6.18 | 1.48 | +4.70 |
| P03 | 9.77 | 0.83 | +8.93 |
| P04 | 5.00 | 5.00 | 0.00 |
| P05 | 13.50 | 17.20 | -3.70 |
| P06 | 8.70 | 1.20 | +7.50 |
| P07 | 6.00 | 0.58 | +5.42 |
| P08 | 6.00 | 8.00 | -2.00 |
| **Mean ± SD** | **7.46 ± 3.02** | **5.31 ± 5.74** | **+2.15 ± 5.07** |
| **Paired t-test** | t(7)=1.20, p=0.27, d=0.42 |  |  |

\*Positive values indicate reduced screen engagement under the AI-scribe condition.

### Screen Time During Consultation

Clinicians spent a mean (±SD) of 7.46 (±3.02) minutes (min) on the screen with traditional documentation versus 5.31 (±5.74) with the AI-scribe, giving a paired mean reduction of –2.15 min (95 % CI –2.10-+6.41). The difference was not statistically significant (paired t(7)=1.20, p=0.27), a result consistent with the small sample (n = 8) and sizeable within-clinician variability (SD of paired differences=5.09). The associated effect size was small–to-moderate (Cohen d=0.42; Hedges g=0.38, 95 % CI –0.28-1.01), suggesting a potential, but unconfirmed benefit. At the individual level, 5 out of 8 participants (62.5%) showed a reduction in screen time when using the AI-scribe, with differences ranging from 4.70-8.93 min. Two participants (25%) experienced increased screen time, and one participant showed no change.

Chi⍰square analyses pairing screen⍰time categories with clinician NASA⍰TLX scores (p=0.243-0.293), CQ items (p=0.293.0.333), and patient⍰experience ratings (p=0.293-0.333) revealed no associations. Nevertheless, shorter AI⍰scribe screen⍰times consistently aligned with higher clinician satisfaction and more favourable patient perceptions, mirroring the direction of the paired⍰sample effects.

### Clinician Questionnaire (CQ)

Under the traditional method, four clinicians (50%; 95% CI 19.3-80.7) reported overall satisfaction, including one who was very satisfied, whereas two expressed dissatisfactions (25%; 95% CI: 4.6-60.0). In contrast, all eight clinicians (100%; 95% CI 68.8-100) described themselves as satisfied with the AI-assisted documentation process, including four who were very satisfied (**Figure 2**). Notably, agreement with the statement “*The documentation method reduced my cognitive load*” increased from 0% (95% CI: 0-31.2) under traditional conditions to 87.5% (95% CI: 52.9-99.4) with the AI-scribe (**Figure 2**). Beyond cognitive relief, clinicians also cited meaningful improvements in consultation flow, sustained attentional focus during interviews, and overall time efficiency. The proportion who judged the documentation process as efficient increased from one of eight participants (12.5%; 95% CI: 0.6-47.1) under the traditional method to eight (100%; 95% CI: 68.8-100) with the AI-scribe; those who considered themselves able to maintain uninterrupted patient focus rose from two (25%; 95% CI: 4.6-60) to eight (100%; 95% CI: 68.8-100). Those who judged the time spent on documentation to be reasonable increased from three clinicians (37.5%; 95% CI: 11.1-71.1) under the traditional method to eight (100%; 95% CI: 68.8-100) with the AI-scribe. Conversely, the proportion who reported feeling rushed declined from five clinicians (62.5%; 95% CI: 24.5-91.5) to 0 (0%; 95% CI: 0.0-36.9).

**Figure 2.**
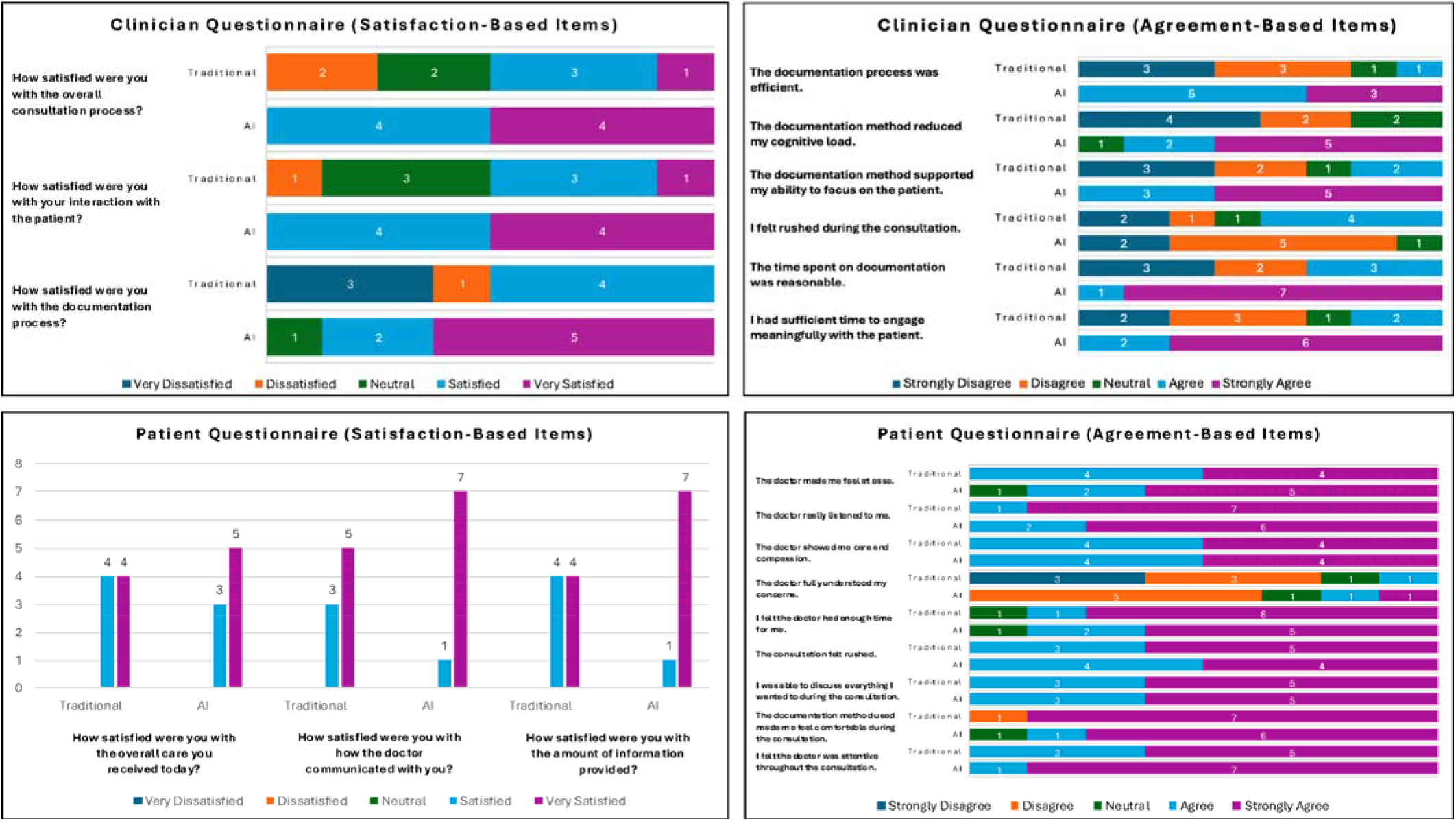
*Clinician and Patient Questionnaire Responses by Documentation Method*. **2A**. Clinician Questionnaire responses to 3 satisfaction-based items, each rated on a 5-point Likert scale (Very Dissatisfied to Very Satisfied). **2B**. Clinician Questionnaire responses to 6 agreement-based items, each rated on a 5-point Likert scale (Strongly Disagree to Strongly Agree). **2C**. Patient Questionnaire responses to 3 satisfaction-based items, each rated on a 5-point Likert scale (Very Dissatisfied to Very Satisfied). **2D**. Patient Questionnaire responses to 9 agreement-based items, presented as stacked bar charts, each rated on a 5-point Likert scale (Strongly Disagree to Strongly Agree) following traditional versus AI-scribe consultations.

### Patient Experience (PEQ)

Under the AI-scribe condition, 62.5%-87.5% of responses per item fell in the “strongly agree” category, reflecting a consistently positive experiential profile **(Figure 2)**. Notably, no standardized patients (vs. three in traditional) reported feeling rushed during AI-assisted sessions.

### Cross-Tabulation Analyses

In the traditional documentation condition, clinicians who rated documentation as efficient were more likely to have actors rate them as attentive (χ^2^(3)=8.0, p=0.046, OR=6.0). All other associations between clinician-reported experience and patient or quality outcomes were non-significant.

In the AI-scribe condition, consultations where clinicians reported not feeling rushed were significantly more likely to be rated higher for both comfort (OR=8.0, p=0.018) and listening (OR=9.0, p=0.023). Agreement by clinicians that documentation time was reasonable was associated with enhanced patient-perceived empathy (OR=9.0, p=0.018). Crucially, clinicians who perceived the AI-scribe as efficient were significantly more likely to produce high quality documentation. This included associations with both the global SAIL score (≥7/10; p=0.018) and total SAIL checklist score (≥22/30; Likelihood Ratio (LR) p=0.032). Additionally, clinicians who agreed that the AI-scribe reduced their cognitive load were more likely to elicit patient ratings indicating the doctor showed compassion and understood their concerns (LR p=0.048). Furthermore, perceived reasonableness of documentation time correlated significantly with inclusion of justified follow-up plans in consultation notes (p=0.005). Notably, clinician satisfaction on the CQ did not correlate significantly with any NASA-TLX domain.

## Discussion

This one of its kind simulation-based study evaluates the impact of an ambient AI scribe on psychiatric documentation in terms of quality, clinician workload, and consultation experience. Compared to traditional keyboard-based methods, we conclude that using AI-scribe significantly reduced clinician-reported workload and frustration, while improving documentation quality as measured by SAIL scores. Clinicians reported greater efficiency, reduced cognitive burden, and enhanced ability to maintain patient focus. On the other hand, simulated patients noted improved empathy, attentiveness, and reduced sense of being rushed. These findings highlight the potential of ambient AI tools to improve clinical consultation dynamics in psychiatry and beyond.

### Clinical Workload

One of the most prominent findings in this study was the significant reduction in clinician workload when using the AI-scribe, particularly in temporal demand, effort, and frustration. These improvements mirror findings from previous studies. For instance, Doshi et al. (2024) reported that ambient AI scribes reduced documentation time and improved workflow satisfaction among oncology providers by offloading administrative tasks without compromising care quality (Doshi et al., 2024). Similarly, Balloch et al. (2024) demonstrated decreased physician burnout and cognitive strain with AI-assisted documentation in primary care settings (Balloch et al., 2024). Our results extend this evidence to the psychiatric context, where documentation demands are especially high due to the narrative depth required in assessments. The dramatic reduction observed in our total NASA-TLX workload scores (mean difference = +436.25 points) emphasise the cognitive relief offered by AI-scribes and supports their role in easing the daily documentation burden in mental health settings.

### Documentation and Clinician Satisfaction

Our study showed an improvement in documentation quality when clinicians used AI-scribe compared to traditional methods. This shifted notes from “Very Poor/Poor” (in the traditional method) – with various missing elements in traditional notes – to the “Fair/Good” band (AI-scribe) and this led to improved satisfaction markedly. This has also been seen previously such as in a recent JAMA study, where using an AI scribe significantly lowered physicians’ cognitive workload (also measured by NASA-TLX) and increased overall work satisfaction, especially among primary care doctors (Stults et al., 2025). Similarly, Duggan et al. reported that an AI scribe reduced time spent on EHR notes by approximately 20% and cut after-hours charting by 30%, yielding “greater efficiency” and a “lower mental burden” of documentation; clinicians also noted enhanced patient engagement (Duggan et al., 2025). In one qualitative report, physicians remarked that scribes let them be more “face-to-face” and build rapport with patients (Shah et al., 2025a). Together, these results suggest that ambient scribes can meaningfully alleviate cognitive strain and allow clinicians to focus on patients. Clinically, this implies that AI scribes could improve workflow efficiency and physician well-being, potentially mitigating burnout and enhancing patient care quality.

### Patient Satisfaction

SPs rated AI-scribe consultations more positively, and interestingly, no SP in our study reported feeling rushed during AI-scribe consultations, suggesting SPs perceived greater satisfaction in terms of attentiveness and empathy when clinicians used the AI-scribe. This also echoes with the findings of Balloch J et al., where parent-actors reported improvement in their experience in the AI scenario compared to traditional (Balloch et al., 2024). In another survey, most patients felt comfortable with remote scribing and reported that it did not undermine their trust in the clinician, even though in this case it was a remote medical scribe via Google Glass face-mounted technology and not a handheld device (Odenheimer et al., 2018). However, qualitative studies have recorded rare instances of patient discomfort: for example, two patients initially consented to AI scribing but later asked their physician to “not use this ever again”, reflecting privacy or connection concerns (Shah et al., 2025a). Even though rare, this contrast hints that while many patients appreciate the added clinician attention afforded by scribes, others may worry about privacy or loss of personal interaction.

Similarly, Shah et al. found that physicians across specialties at Stanford Health Care valued AI scribes for enabling reduced task load and for minimizing burnout due to documentation, benefits that closely mirror the psychiatric clinicians’ appreciation of improved focus on the patient since the documentation is taken care of by the AI scribe (Shah et al., 2025b).

From the patient perspective, Moy et al. concluded that most of the patient were comfortable with AI-assisted documentation in consultations for cancer care, neurosurgery, preventive medicine, radiology, and virtual care among others, along with a concern towards privacy policies, informed consent, and patient understanding of how the AI scribe will be used (Moy et al., 2024). Another study noted that patients expressed ‘curiosity instead of concern’ and may have felt “different” while sharing sensitive information during AI consultations, which may affect the information exchanged during those patient-clinician interactions (Evans et al., 2025). This denotes that patients might require additional information about data storage and security to make an informed decision. Collectively, these findings support the integration of ambient AI scribes but highlight the need for thoughtful implementation strategies that preserve the trust, empathy, and relational nuance central to psychiatric care.

### Strengths and Limitations

As the first to assess the use of an ambient AI scribe in psychiatric consultations, this study used a rigorous simulation-based, crossover design in which each clinician acted as their own control, minimizing inter-participant variability. Blinded documentation assessment and complete data collection further strengthened internal validity. However, certain limitations must be acknowledged.

The present study is exploratory and was conducted with a small sample of eight clinicians from a single centre, limiting both statistical power and external generalisability. Although simulation offers a controlled environment, it may not fully capture the complexities and relational nuances of real-world clinical encounters. Additionally, with the natural learning curve, the subsequent encounters might be anticipated by the participating physicians, thus it may have skewed the results.

## Conclusions

This is the first study to assess the utility of ambient AI scribes among psychiatrists, demonstrating improved documentation quality, reduced workload and enhanced clinician and patient consultation experience. These findings suggest the growing potential for AI scribes as a tool for enhancing patient care and combatting clinician burnout.

## Data Availability

Data collected and analysed , including an research materials, will be readily shared upon direct request by email to the corresponding author of this study.

## Acknowledgements

The research team acknowledges the support of Mohammed Bin Rashid University of Medicine and Health Sciences in allowing the study to be conducted at their simulation center.

## Data Access, Responsibility, and Analysis

The principal investigator (FAN), co-investigators (SAB and RK) had full access to all the data in the study and take responsibility for the integrity of the data and the accuracy of the data analysis.

## Data Sharing Statement

Data collected and analysed, including an research materials, will be readily shared upon direct request by email to the corresponding author of this study.

## Transparency Declaration

The lead author and manuscript guarantor affirm that the manuscript is an honest, accurate, and transparent account of the study being reported; that no important aspects of the study have been omitted.

## Author Contribution

Faisal A. Nawaz contributed to conceptualization, methodology, writing original draft, writing review and editing, supervision, and project administration. Syed Ali Bokhari contributed to conceptualization, methodology, writing original draft, and writing review and editing. Firdous M. Usman contributed to conceptualization, methodology, data curation, and writing review and editing. Zara Arshad contributed to writing original draft and writing review and editing. Meghana Sudhir contributed to methodology, validation, and writing review and editing. Ralf Krage contributed to methodology, validation, and writing review and editing. Rahul Kashyap contributed to conceptualization, methodology, and writing review and editing.

