## Supplemental Figure for "Evaluating an Ambient Artificial Intelligence Scribe for Documentation Quality and Efficiency in Psychiatric Consultations: A Simulation-based Study"

**eTable 1**. *Study outcome measures with their classification, data source, and rationale for inclusion*.

| **Outcome** | **Type** | **Source** | **Justification** |
| --- | --- | --- | --- |
| *National Aeronautics and Space Administration Task Load Index (NASA-TLX) total score* ^25^ | Primary | Clinician self-report | Widely validated tool assessing cognitive and physical workload |
| *Sheffield Assessment Instrument for Letters (SAIL) total score* ^17^ | Secondary | Independent rater | Standardised and validated assessment of outpatient documentation |
| *Clinician Questionnaire (CQ)* | Secondary | Clinician self-report | Captures perceived usability, satisfaction, and consultation flow |
| *Patient Experience Questionnaire (PEQ)* | Secondary | Simulated patient actor | Proxy for real-world patient-centred interaction and satisfaction |
| *Documentation time* | Exploratory | Observer stopwatch | Objective measure of documentation burden |
| *Eye contact duration* | Exploratory | Video analysis | Proxy for relational engagement and clinician–patient connection |
